# Cluster analysis of epidemiological characteristic features of confirmed cases with the novel coronavirus (COVID-19) outside China: a descriptive study

**DOI:** 10.1101/2020.06.28.20142000

**Authors:** Minh-Duc Nguyen-Tran, Ali Ahmed-Fouad Abozaid, Luu Lam Thang Tai, Le Huu Nhat Minh, Quang-Loc Le, Hoang-Dung Nguyen, Khanh-Linh Dao, Huu-Hoai Le, Nguyen Tien Huy

## Abstract

**Background:** Novel coronavirus COVID-19 has caused significant global outbreaks outside China. Many countries have closed their borders with China and performed obligate protective procedures, however, this disease was still rising worldwide. In this report, we aim to identify transmission patterns from China to other countries, along with describing the disease control situation of countries.

**Methods:** We retrospectively collected information about infected cases with COVID-19 from WHO situation reports, official notification websites of health ministries and reliable local newspapers from each country. Descriptive and cluster analysis was performed to describe the transmission characteristics while the logistic regression test was used to estimate the risk factors for the occurrence of an infected individual with an unknown source.

**Results:** A total of 446 infected cases were recorded from 24 countries outside China until 12 February 2020, with the number of reported infected cases were doubled every 3.08 ± 0.35 days (range from 2.6 to 3.9). Besides the spread from China, the transmission was originated from sub-endemic countries (Japan, Thailand, Singapore, Malaysia, France, German). Out of 6 countries got occurrence of an infected individual with unknown source and possible potential factors contributed to this occurrence was a time of epidemic circulating, number of patients and number of clusters when the occurrence still has not happened, and notably, the unreported situation of Chinese tourists’ information.

**Conclusions:** The situational reports of each country about COVID-19 should be more detailed mentioning the transmissions routes with keeping contact tracing of the unknown cases to increase the control of this disease.

## BACKGROUND

Novel coronavirus is a new strain of the previously discovered Coronaviridae family, a member of RNA viruses of the order Nidovirales which own spiky projections on their outer surfaces that resemble the shape of a crown, or “corona” in Latin^1^. They are zoonotic viruses infecting mainly birds and mammals, in addition to humans. Human coronavirus infections are mostly mild, however, in the past two decades, two beta-coronaviruses; the Severe Acute Respiratory Syndrome Coronavirus (SARS-CoV), and the Middle East Respiratory Syndrome Coronavirus (MERS-CoV) have infected more than 10,000 patients over a potentially short period of time^1-5^. With mortality rates of 10% for SARS-CoV and 37% for MERS-CoV, respectively, they are one of the most devastating epidemics all over the world^6^.

On December 31, 2019, an outbreak of severe acute pneumonia with unknown etiology occurred in Wuhan with people who attended a wild animal and seafood market in China. China discovered at this time a new strain of coronavirus resembles SARS-CoV and has informed the world health organization (WHO) about this new catastrophe and new countries have taken obligate procedures to prevent the virus from spread abroad^7,8^. However, Multiple countries of almost all continents including the USA, Vietnam, Germany and Australia have reported confirmed cases of SARS-CoV-2^9^. COVID-19 possesses an exceptional ability of transmission and no specific treatment for it has been reported. Infected patients are managed with supportive care, but antiviral medications and combination treatment have been reported to be used^10^. The clinical symptoms of SARS-CoV-2 have been reported as fever, dyspnea, cough, abdominal distress, and in severe cases, it could lead to complications such as acute respiratory distress syndrome (ARDS), pulmonary edema and multiorgan failure^11^.

Furthermore, efforts in measures implemented so far have been directed towards preventing and containing the risk of spread beyond such local clusters to prevent extensive community spread. Recently, an asymptomatic transmission has been reported in many countries including Germany, South Korea, China^12-14^; WHO has reported that the transmission occurred easily by face to face through coughing, sneezing, contact with secretions of infected people like nasal and oral droplets, tears or substantially by fecal-oral route^15^. The basic reproduction rate of COVID-19 estimated at 3.28 representing the highly contagious transmissibility of this disease putting us on a great challenge for containment this largely infectious disease outside China^16^. In this report, we aimed to investigate the epidemiological importance of clustering the COVID-19 confirmed cases outside China to increase the awareness of these countries to develop more appropriate surveillance system ways to limit the spread of SARS-CoV-2 and contact trace of their unknown cases.

## METHODS

### Data collection and search strategy

We initially obtained data from a manual search of clinically confirmed COVID-19 through ProMED, WHO, CDC, ministries of health of different countries press releases, journal case reports like Lancet and New England Journal of Medicine (NEJM) and local news. We used since then many search keywords to get these data including ‘novel coronavirus’, ‘COVID-19’, ‘SARS-CoV-2’, ‘2019-nCoV’, ‘cluster’, ‘transmission’ and ‘contact tracing’ ‘Wuhan’ and others. The data acquired have been sorted according to demographic characteristics such as age, sex, country, history of contact or travel, announcement date and the transmission clusters. February 12, 2020 was the last date we have done these searches.

### Sources reliability

We have measured the reliability of our sources using The Journal of the American Medical Association (JAMA) four benchmarks published in 1997. The four criteria involved the authorship (the information of authors or writers and their affiliation should be provided), attribution (the sources have to list their references prominently), disclosure (ownership, funding source, advertisement and conflict of interests should be obvious for readers) and currency (the date of publishing and updating contents should be posted)^17^.

### Ethical condition

All data have been acquired from online sources and governments’ websites without any direct contact data from patients.

### Definition of epidemiology of infectious clusters and doubling time

We defined the transmission clusters epidemiologically into four main types, A, B, C, and D, based on the infectious source and risk of an outbreak. A cluster was classified into group A if the primary pathogen had a history of traveling to China or have been in contact with foreigners abroad outside countries, who were later confirmed SARS-CoV-2-infection. Because it was impossible to determine who exactly spread the disease in the case of an infected group returned from China, each of the patients in the group would be considered as an independent primary pathogen and established an independent cluster. Group B involved clusters that patients without traveling to China but got local contact with Chinese tourists. Due to the information of Chinese tourists was not disclosed and such tourists were not reported as infected cases, we considered clusters in group B to be at higher risk of an outbreak than group A. Groups C and D, at more risk, included clusters where the source of infection could not be identified. While patients in a cluster C were involved with a specific area, such as a church, a conference, etc., information of patients in clusters D remained completely unknown.

The doubling time in our study is defined as the time required for the number of reported cases to double. It was calculated based on the formula: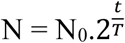. Because we only could record the date of diagnosis (not the date of expose), the value hence only partially reflected the rate of disease transmission.

### Statistical analysis

Descriptive demographic and cluster analysis have been conducted inclusively for this dataset. The missing data have been considered as unreported due to some governments didn’t reveal these data to protect the personality of their patients. Logistic regression then was performed to determine relationships between potential factors and the occurrence of an infected with unknown source, with the significant value chosen was < 0.05.

## RESULTS

### Demographic characteristic of infected patients

We have identified 446 cases from 24 countries confirmed with COVID-19 infection outside China. The median (IQR) number of reported cases doubling time was 3.08 ± 0.35 (range from 2.6 to 3.9) days and recorded the highest increase of reports (n = 113) between 10-Feb and 11-Feb 2020, mostly from cases of Diamond Princess (**Figure 1)**^18^. Concisely, the most affected country with the biggest number of infected patients was Singapore with a total number of 50 since the beginning of the outbreak up to February 12, 2020 (**Table 1)**^19^. The first case outside China was reported in Thailand on January 13, 2020^20^. Further details of the sequences of these cases in each country are presented in the ***Supplementary document***.

**Table 1:**
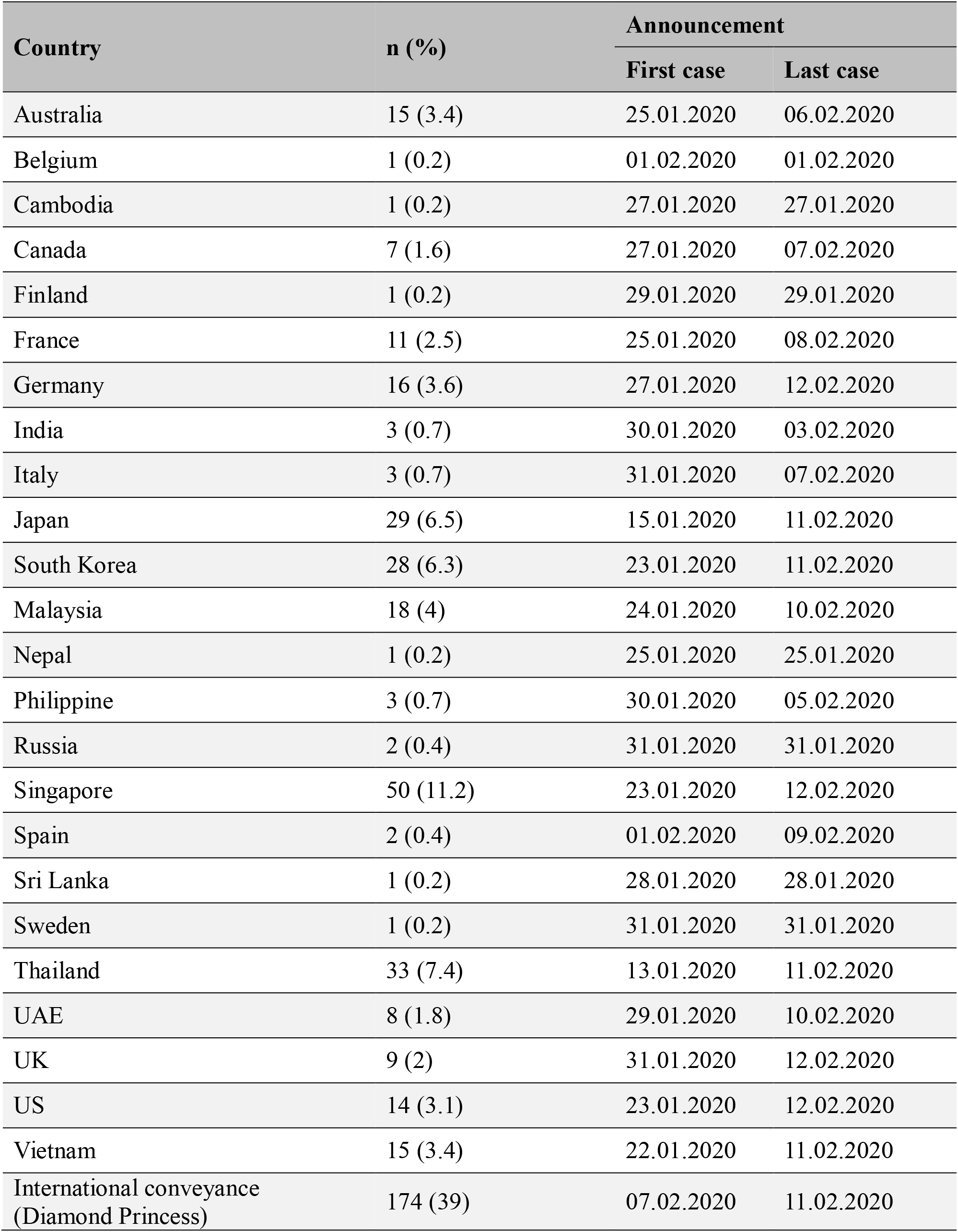
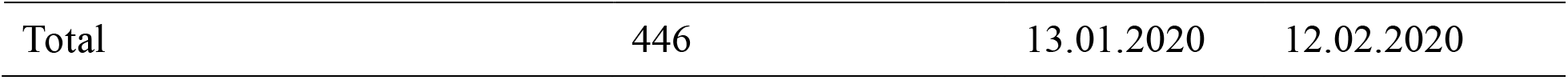
Summary of infected cases around the world outside China (n = 446)

**Figure 1:**
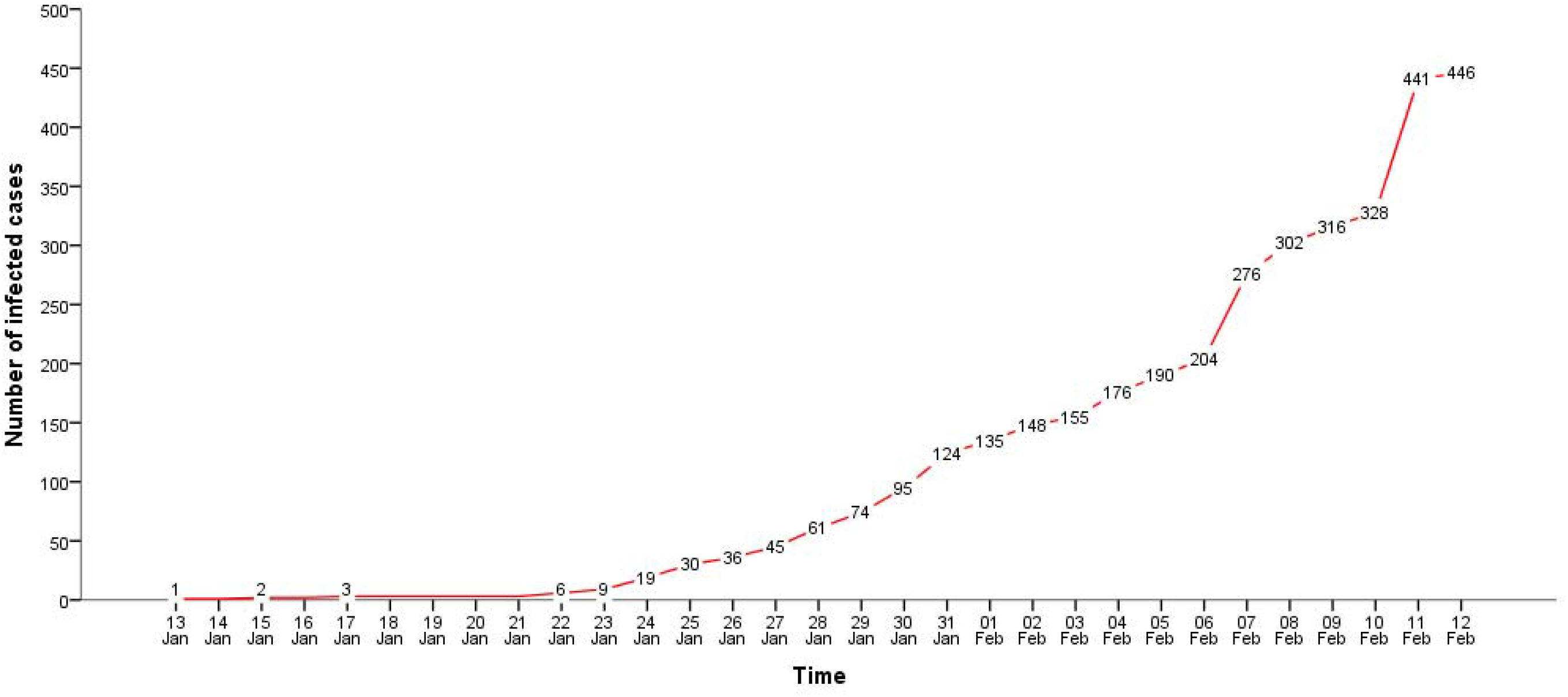
The total number of infected patients globally since the first case was reported until February 12, 2020. **Legend:** Doubling time 3.08 ± 0.35 days

Approximately, 57% of the COVID-19 infected patients were males compared to 43% female patients. The median (IQR) age was 42 (32-56), and the youngest infected patient was a three-month old child from Vietnam^21^. All patients survived up to this date (February 12, 2020), and some were even discharged from the hospital except for one case who died on February 2, 2020. The patient was a 44-year old male passenger coming from Wuhan to Philippines^22^. Other demographic characteristics of the included patients were presented in **Table 2**.

**Table 2:**
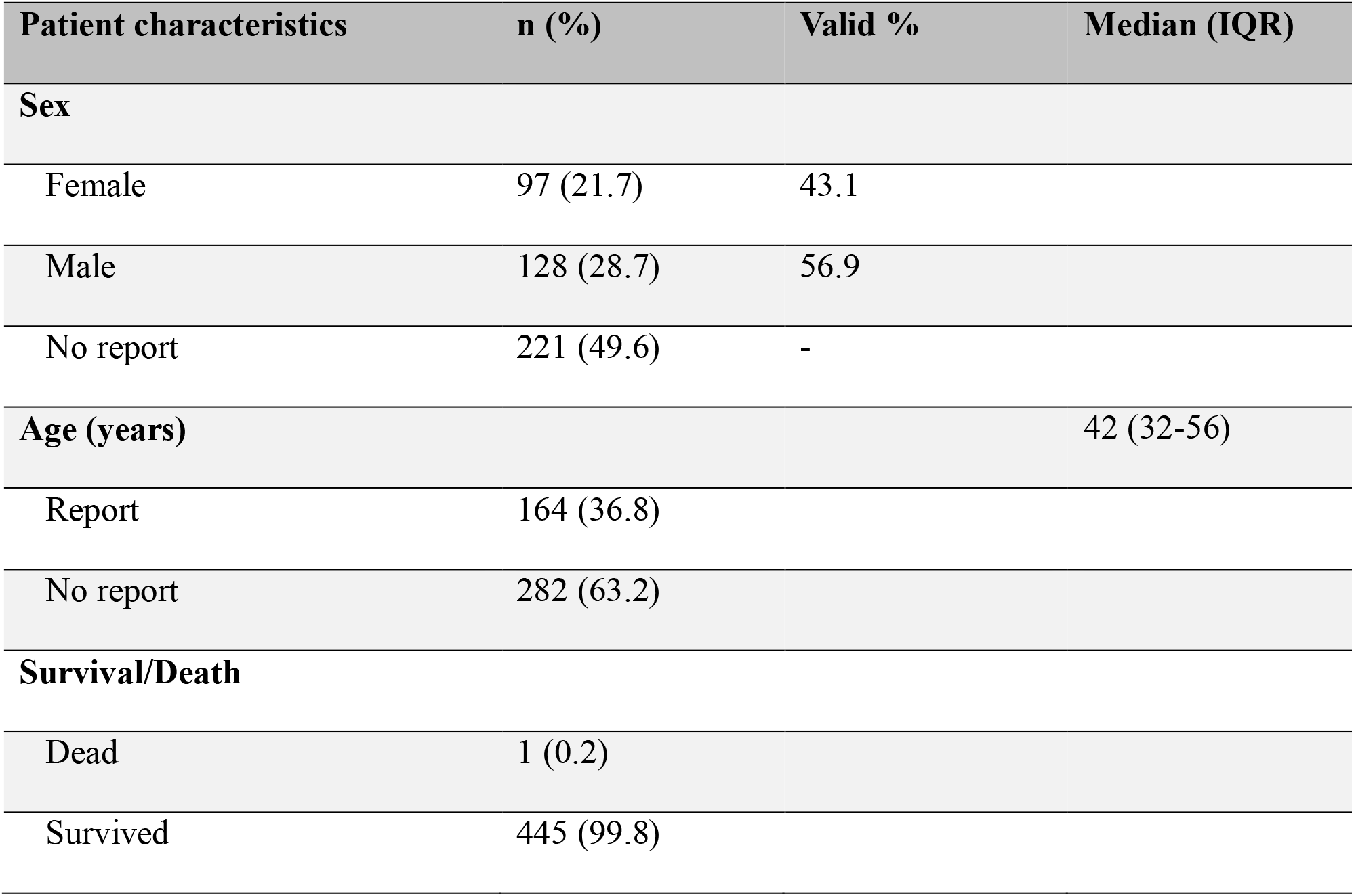
Demographic features of the infected cases (n = 446)

### Transmission outside China

A total of 214 transmission clusters from 446 patients were determined. There were 178 clusters (83.2%) that were involved with China and 12 clusters (5.6%) were spread from other sub-endemic countries (Japan, Thailand, Singapore, Malaysia, France, German). Spain was the only country in which the spread of COVID-19 from China was completely indirect, and Korea represented the most counties have indirect transmission. More details of these transmissions were shown in **Figure 2**.

**Figure 2.**
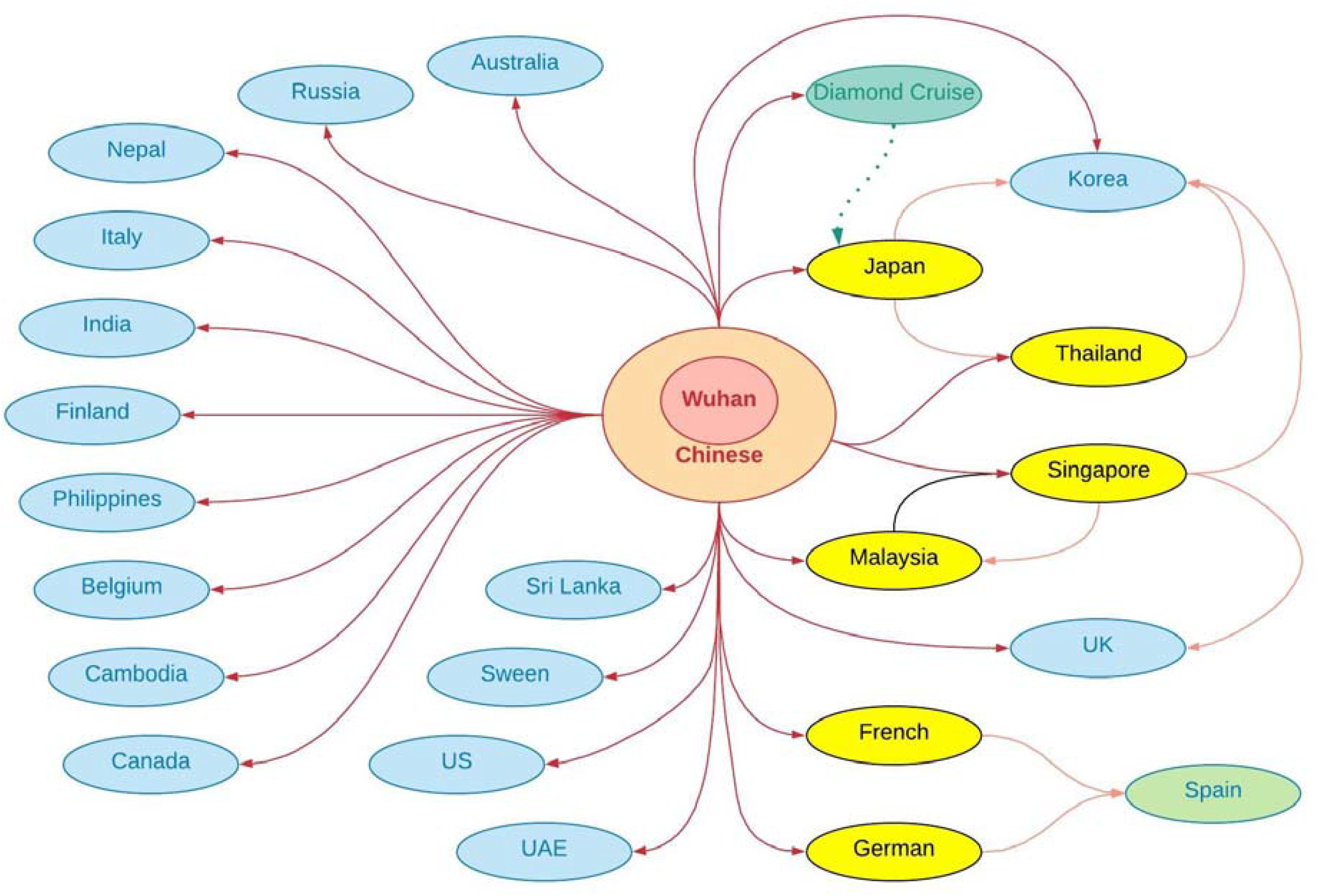
The transmission of COVID-19 from China to other countries.

**Table 3** showed a summary of the size and the type of clusters. The biggest cluster recognized was the Diamond Cruise ship, with a total of 175 infected patients^18,24-26^, followed by the cluster in Germany (13 patients), Singapore (9 patients), France, Viet Nam, UK, Korea (6 patients) ^19,21,27^. Notably, Results also have shown that the number of clusters of Group A, B, C and D was respectively 182 (85.0%), 9 (4.2%), 10 (4.7%), 13 (6.1%). When approaching group, A, B, we found that 168 non-transmissible clusters (1-patient cluster), 18 clusters confirmed the local transmission between primary pathogen and local people, and 5 clusters recognized the transmission between local people.

**Table 3:**
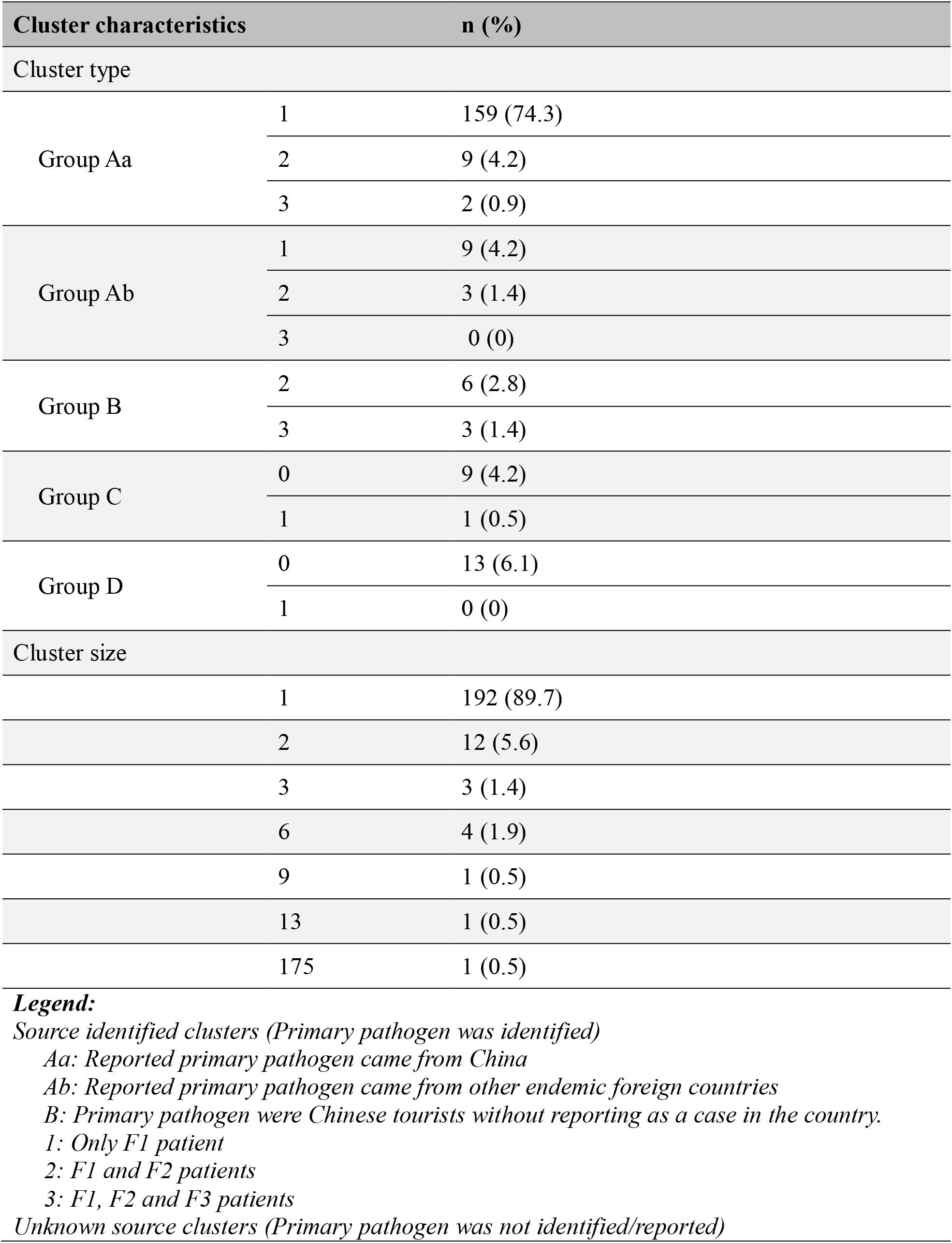
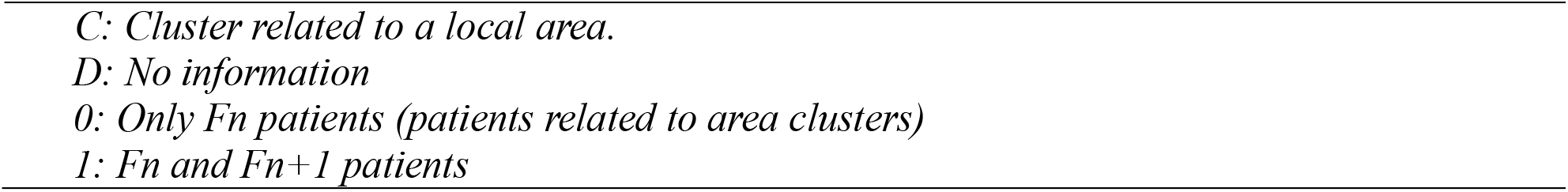
Demographic features of the transmission clusters (n = 214)

At the national level, out of 24 countries infected with COVID-19, the occurrence of an infected individual with unknown source happened in 6 countries (Australia, Germany, Japan, Singapore, Thailand, UAE). Summarily, there were 13 countries that local human-human transmission began, with the number of local transmission (F2 and F3) range from 5.6% in Malaysia to 55.6% in UK from identified primary pathogen (F1). Other information has been summarized in **Table 4**.

**Table 4:**
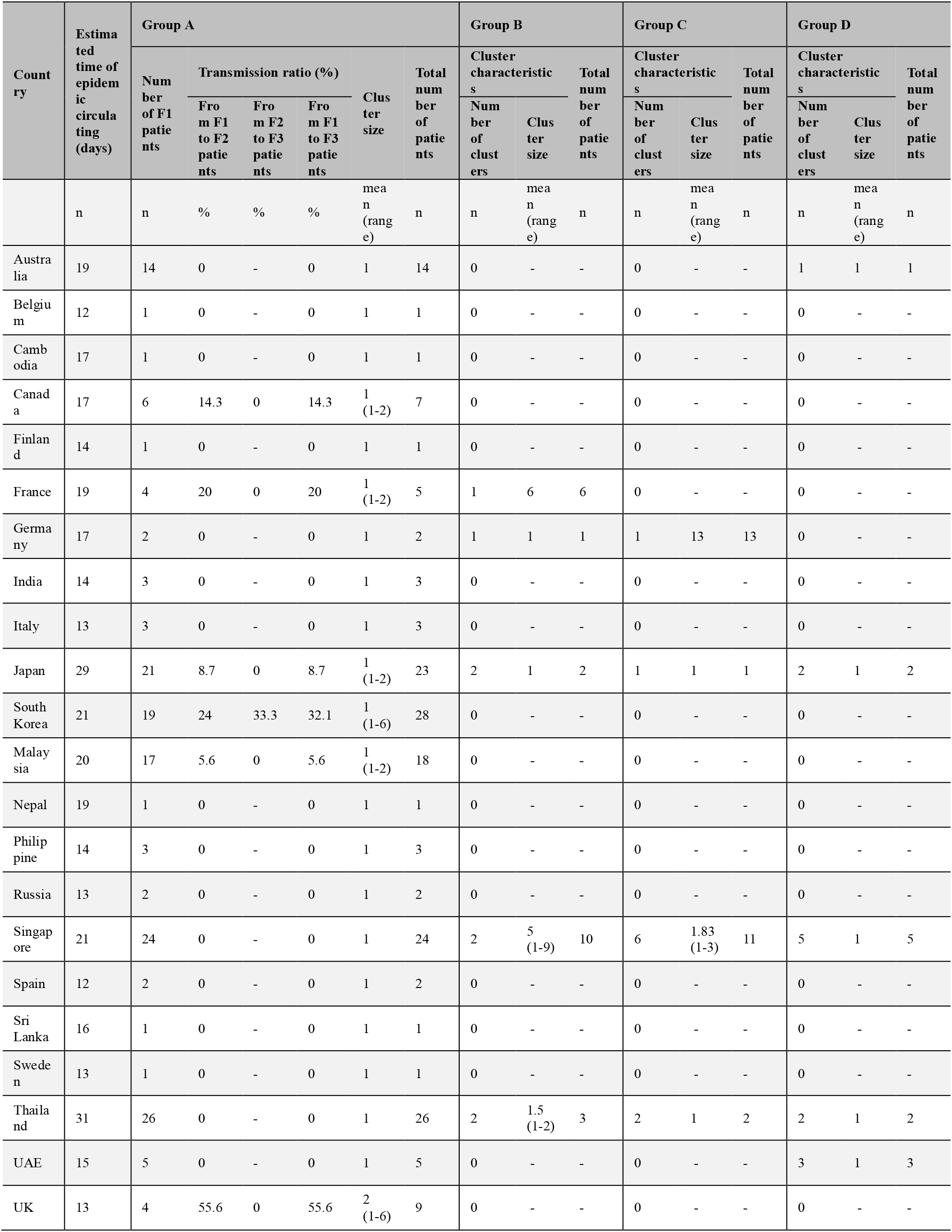
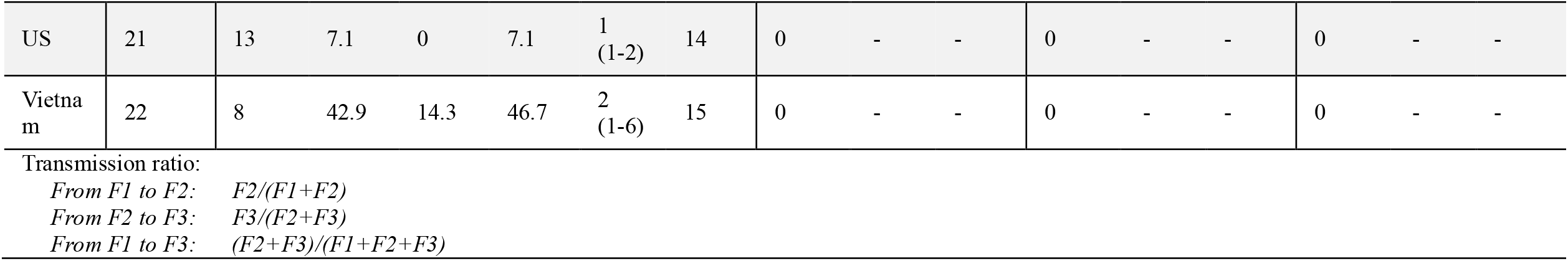
Epidemiology of the transmission clusters across countries.

### Occurrence of an infected individual with unknown source

**Table 5** showed the associated factor with the occurrence of an infected individual with unknown source. Time period from the first case reported until Feb 12 was related with increasing of the risk (p = 0.044) (OR: 0.76) [95% CI: 0.58-0.99]. Notably, countries which was unreported information of Chinese tourists was associated with highly significant risk of occurrence event (p = 0.001). We also detected other factors that can contribute on the situations like the number of control patients (p = 0.032) (OR: 0.89) [95%CI: 0.81-0.99] and number of control clusters (p = 0.017) (OR: 0.85) [95%CI: 0.74-0.97].

**Table 5:**
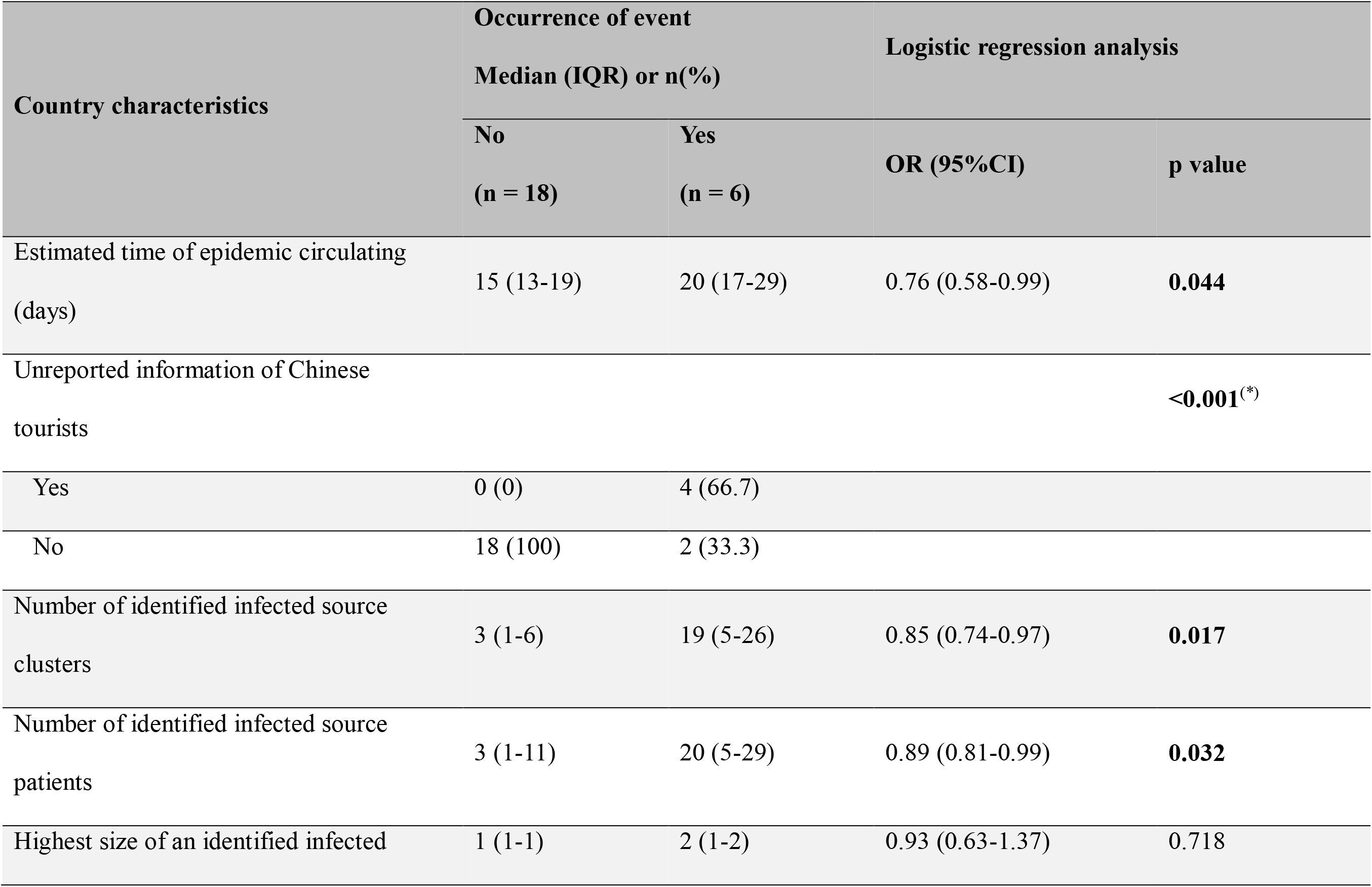
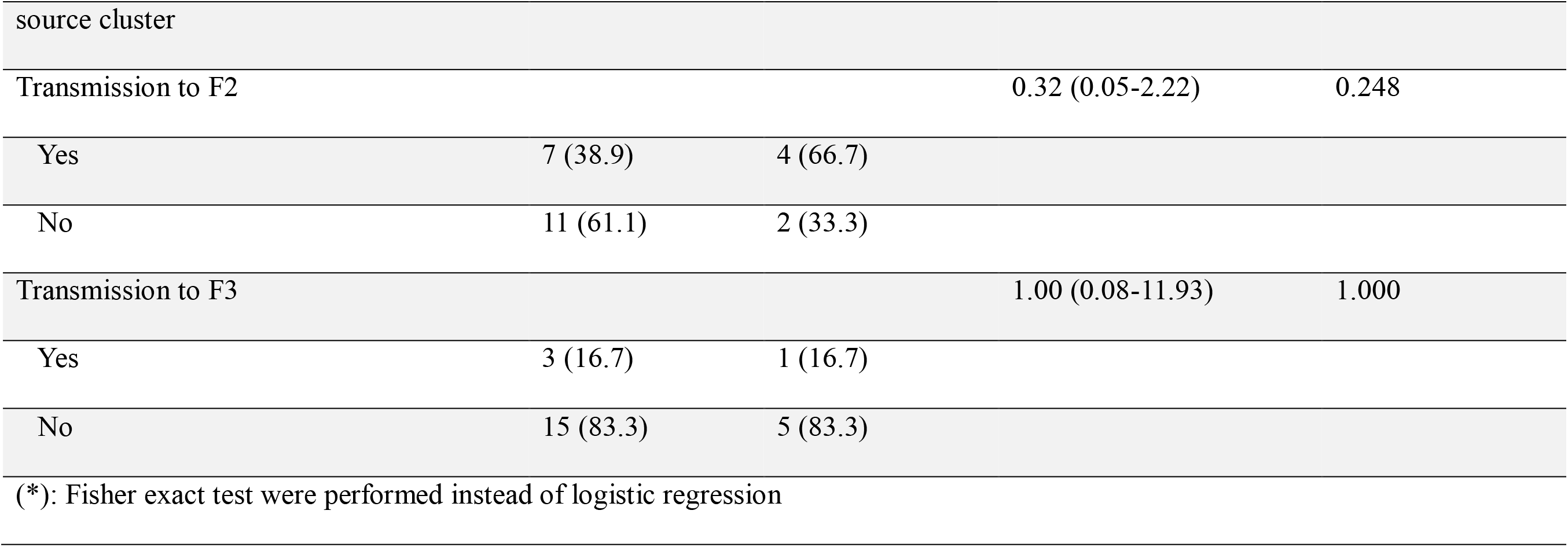
Factors associated with occurrence of an infected individual with unknown source (N = 24)

## DISCUSSION

The overview of our study has undoubtedly affirmed that the number of patients diagnosed with the newly discovered coronavirus has continued to increase every day, affecting many countries all over the world. This virus unfortunately showed a very interactive easily transportation from person to person. Liu et. al. has detected the mean reproductive rate (R0) of severe acute respiratory syndrome coronavirus 2 (SARS-CoV-2) estimated around 3.28 compared to 3 in SARS^28,29^. However, some studies have been gone far and estimated R0 of this new virus to reach above 6 in average indicating the hard situation of the world for the inability to control this emergent disease^30,31^. Therefore, many countries have put restrictive measures on travelers coming from China to limit the spread of the disease among their populations. In China, the authorities decided to lock down many cities to control the disease which caused people of different nationalities to seek their home return. As a result, the epidemiology and transmission of the disease are vague concerning any possible transmission by asymptomatic patients as the incubation period of COVID-19 is 14 days. On the other hand, a study reported the doubling time of the outbreak inside China to be 7.4 days on January 22, 2020^6^. The mean doubling time we calculated here was every 3.08 ± 0.35 days outside China (**Figure 1**). However, we could not compare both estimates to each other, and other similar outbreaks due to lack of sufficient data of expose time in our sources unlike they had. Therefore, we couldn‘t either assess R0, besides, further investigations are needed regarding this concern as this rate was likely to be increased with time.

Currently in this study we have attempted for more investigation and clustering the epidemiology of the population from outside China who have been related to the outbreak until February 12, 2020. To this date, 446 cases were reported internationally in 24 countries with the highest number of patients among the Singaporean population, and the lowest in Belgium^32^, Cambodia^33^, Finland^34^, Nepal^35^, Sri Lanka^36.^and Sweden^37^ which reported only one case each. The huge number of infected patients is due to the Diamond Princess cruise (n = 174) which was locked down on February 3, by the Japanese authorities after COVID-19 was positively confirmed in Hong Kong Chinese man ^24-27^. Without these cases, Singapore would have the largest record of on-land confirmed cases of COVID-19 (n = 50)^19^ **(Table 1)**. The primary transmission (Group A) represented the majority that brought the primary pathogen (n = 168). Moreover, we did more subgrouping of these cases to know where most of cases mostly came from and found the majority of them are Chinese people who recently returned from China (Aa) (n = 159), however, some cases recognized from other sub-endemic countries (Ab) (n = 9) as in Japan^38^, Singapore^39^, France^40^, Germany^41^, Thailand^42^ and Malaysia^43^. highlighting that the governments should not only focus on checking people from China, but also, in countries that reported COVID-19 infected cases. In addition, we noticed some of the reported cases had no history of travel to China, where the outbreak took place but had been in close contact with Chinese people, suggesting person to person transmission (F2) which is similar to the (SARS)^44^ and (MERS)^45^ outbreaks in 2003 and 2013, respectively **(Table 3, Figure 2)**.

The primary cases who infected other persons later (F1) were greatest in Thailand (n = 26), Singapore (n = 24), and Japan (n = 19) which is suggestive of the poor tracking the patients entering the country **(Table 4)**. In Thailand, a taxi driver was reported to have been infected on January 28, 2020. The patient had no history of travel to China but was in contact with a Chinese tourist which the first case of a person to person transmission in the country^46^. Moreover, transmission with difficulty in detecting the source of infection, and infections with no detectable source (Group C and D) (either not reported or cannot be detected) was presented in some countries. In Australia, UAE and Germany reported 1, 3 and 1 respectively in these two groups, while Asia recorded the highest number of such cases reaching 11 cases in Singapore, 3 in Japan (**Table 4)**. These are serious numbers and can be misleading as suggestive of the emergence of the disease in these counties with no transmission from China being reported. However, they were estimated in the early period, so they were not always reflected the current statements as some country may gain the ability to find the links of their cases like Singapore latterly but still give some predictions of future events in other countries like what happened later in Japan and Germany. To note, we try to develop a new way of predictions and rapid judgmental tools of ongoing daily situations report for those countries who care to identify and clarify their present conditions. Therefore, authorities should exert more effort in reporting all cases in an evidence-based manner. The earliest case to be reported outside China was in Thailand on January 13, 2020 who had been to China lately^20^. Besides, the only patient that died from COVID-19 from outside China was reported in Philippine on February 2, 2020^22^. However, the Death numbers were likely to be increased in the following days because of the continuously increasing numbers of confirmed infected patients.

The pathophysiology of this COVID-19 has not powerfully known, but Sohrabi et. al. said that most of the transmission occurred in the symptomatic period with increasing some reporting of asymptomatic transferring cases^47^. Subsequently human to human transmission was reported in Vietnam^48^, Japan^49^ and USA^50^. Some cases are asymptomatic^12,51,52^ due to the 2-week incubation period of the virus which makes it hard to detect these cases, further, possible transmission of the virus to other contacts is easy. In addition, a Vietnamese case was reported with normal clinical findings and mild symptoms^53^. We couldn‘t report these findings due to non-adequate information in the majority of cases. Nonetheless, we kept tracked to know about the transferring patterns of this disease in different countries, then we found the direct transmission ratio from Chinese people (F1 to F2) has recorded the highest in the UK (55.6%), Vietnam (44.6%) and Republic of South Korea (24%) while the indirect local ratio (F2 to F3) revealed in South Korea (33.3%) and Vietnam (14.3%) can represent a hazard ratio of outbreak occurrence but still remained non-significant (p > 0.05) (**Table 4, Table 5**). However, the impact of unreported Chinese cases in out of control countries was a significantly high risk of the outbreak (p < 0.001). The time of announcement of cases and the number of control patients or clusters are still risk factors to produce pandemic disease (p < 0.05) **(Table 5)**. The cluster size didn‘t show a significant this risk attribution (p > 0.05), in spite of that, this assessment has been made in the early stage when the super-spreader factors haven‘t grown yet. Consequently, healthcare officials must put restrictive measures at all levels for rapid detection of the virus on people coming from China or those countries which became lastly an epicenter of this disease.

Eventually, we would like to recommend the diagnostic measures of the COVID-19 to be more developed. Mostly we used RT-PCR techniques according to the WHO recommendations^54^.

Samples were effectively positive for the virus from the upper and lower respiratory tract^6^, and stool^55^. Furthermore, the virus was detected in the serum of severely ill patients in China^56^.

However, Ai et. al. had another opinion for considering the chest computed tomography (CT) as a primary tool for investigation^57^. He foresaw that the CT scan was much easier and more practical. Another study suggested a combination of both techniques when RT-PCR results were negative but it should be in highly suspicious persons ^58^. We also agreed on prevention of the outbreak should include suspected and confirmed cases isolation, rapid monitoring to the contacts with these cases, and preparing the healthcare facilities and training the medical stuffs for dealing with such cases. Health education of people who have been in close contact with infected patients for rapid presentation at nosocomial procedures is crucial in controlling the outbreak. Besides, advising susceptible individuals about the importance of hygiene and wearing personal protective equipment (PPE), and staying away from infected or suspected patients is important to prevent person-to-person transmission^59,60^.

While providing more insight into how COVID-19 spread worldwide, highlighting the importance of coordination between clinicians and public health authorities at the local, state, and federal levels, as well as the need for rapid assessment of clinical information related to the care of patients with this emerging infection, it was undoubtedly that there still were several limitations to our study. Incomplete data especially expose time and incubated period of the infected cases, as most of the reports were published as news reports, Ministries of Health press releases with a short description. Furthermore, investigation hence was needed to gather more knowledge about this disease.

## CONCLUSION

In this study, we reported the epidemiological features of the COVID-19 outside China. Further investigations for developing proper Identification of SARS-CoV-2 cases through daily national situations reports with keeping contact tracing for unknown patients is crucial to maintain the status of countries on control until finding the best cure for this disease.

## Data Availability

All data generated or analysed during this study are included in this published article [and its supplementary information files].

## LIST OF ABBREVIATIONS

“SARS-CoV”: Severe Acute Respiratory Syndrome Coronavirus.
“MERS-CoV”: Middle East Respiratory Syndrome Coronavirus.
“SARS-CoV-2”: Severe Acute Respiratory Syndrome Coronavirus 2.
“WHO”: World Health Organization.
“ARDS”: Acute Respiratory Distress Syndrome
“CDC”: Centers For Disease Control And Prevention.
“ProMed”: An online news site for international society for infectious diseases.
“USA”: United States of America.
“UK”: United Kingdom.
“UAE”: United Arab Emirates.
“PPE”: Personal protective equipment.
“NEJM”: The New England Journal of Medicine Journal
“JAMA”: The Journal of the American Medical Association

## DECLARATIONS

### Ethics approval and consent to participate

Not applicable.

### Consent for publication

Not applicable.

### Authors’ contributions

MDNT was responsible for the idea, AAFA and NTH were responsible for the study design. MDNT, AAFA, LHNM, HDN, KLD and HHL have helped in the collection of dataset. HDN contributed in the drawing of the diagrams of the cases in supplementary material. Data analysis, tables and figures were made by LLTT. MDNT, LLTT, AAFA and NTH interpreted the data results. QLL, LLTT, AAFA and LHNM have contributed in the writing. Final review done by NHT, LLTT and AAFA. The final approval of this version have been made by all participated authors.

## Acknowledgments

None.

